# Association of the use of hearing aids with the conversion from mild cognitive impairment to dementia and progression of dementia: a longitudinal retrospective study

**DOI:** 10.1101/2020.01.31.19015503

**Authors:** Magda Bucholc, Paula L. McClean, Sarah Bauermeister, Stephen Todd, Xuemei Ding, Qinyong Ye, Desheng Wang, Wei Huang, Liam P. Maguire

**Affiliations:** Cognitive Analytics Research Lab, School of Computing, Engineering & Intelligent Systems, Ulster University, BT48 7JL, United Kingdom; Northern Ireland Centre for Stratified Medicine, Biomedical Sciences Research Institute, Ulster University, BT47 6SB, United Kingdom; Department of Psychiatry, Warneford Hospital, University of Oxford, OX3 7JX, United Kingdom; Altnagelvin Area Hospital, Western Health and Social Care Trust, BT47 6SB, United Kingdom; Fujian Provincial Engineering Technology Research Centre for Public Service Big Data Mining and Application, College of Mathematics and Informatics, Fujian Normal University, Fuzhou, Fujian, 350108, China; Department of Neurology, Fujian Medical University Union Hospital, Fujian, Fuzhou 350001, China; Department of Otolaryngology, Fujian Medical University Union Hospital, Fujian, Fuzhou 350001, China

**Keywords:** dementia, Alzheimer’s disease, mild cognitive impairment, MCI, dementia onset, dementia incidence, hearing impairment, hearing loss, hearing aid, risk factor, disease progression, cognitive decline, survival analysis, National Alzheimer’s Coordinating Center

## Abstract

**INTRODUCTION:** Hearing aid usage has been linked to improvements in cognition, communication, and socialization, but the extent to which it can affect the incidence and progression of dementia is unknown. Such research is vital given the high prevalence of dementia and hearing impairment in older adults, and the fact that both conditions often coexist. In this study, we examined for the first time the effect of the use of hearing aids on the conversion from mild cognitive impairment (MCI) to dementia and progression of dementia.

**METHODS:** We used a large referral-based cohort of 2114 hearing-impaired patients obtained from the National Alzheimer’s Coordinating Center. Survival analyses using multivariable Cox proportional hazards regression model and weighted Cox regression model with censored data were performed to assess the effect of hearing aid use on the risk of conversion from MCI to dementia and risk of death in hearing-impaired participants. Disease progression was assessed with CDR® Dementia Staging Instrument Sum of Boxes (CDRSB) scores. Three types of sensitivity analyses were performed to validate the robustness of the results.

**RESULTS:** MCI participants that used hearing aids were at significantly lower risk of developing all-cause dementia compared to those not using hearing aids (hazard ratio [HR] 0.73, 95%CI, 0.61-0.89; false discovery rate [FDR] *P*=0.004). The mean annual rate of change (standard deviation) in CDRSB scores for hearing aid users with MCI was 1.3 (1.45) points and significantly lower than for individuals not wearing hearing aids with a 1.7 (1.95) point increase in CDRSB per year (*P*=0.02). No association between hearing aid use and risk of death was observed. Our findings were robust subject to sensitivity analyses.

**DISCUSSION:** Among hearing-impaired adults, hearing aid use was independently associated with reduced dementia risk. The causality between hearing aid use and incident dementia should be further tested.

**Highlights:**

- High prevalence of dementia and hearing impairment in older adults
- Hearing aid (HA) use associated with a lower risk of incident dementia
- Slower cognitive decline in users than non-users of HA with mild cognitive impairment
- The relationship between hearing impairment and dementia should be further tested

## 1 Background

The escalating costs and devastating psychological and emotional impact of dementia on affected individuals, their families and caregivers makes the prevention, diagnosis, and treatment of dementia a national public health priority worldwide [1]. While the process of drug development to delay the onset of dementia has been slower than initially hoped, there is evidence that behavioural and lifestyle interventions might reduce dementia risk [2-4]. Numerous studies have investigated the effects of physical exercise, healthy diet, and management of medical conditions, such as diabetes and heart disease, on cognitive decline and risk of developing dementia [2,5]. However, there is a paucity of research on hearing impairment and dementia. Such research is vital, given the high prevalence of dementia and hearing impairment in older adults, and the fact that both conditions often coexist.

The prevalence of hearing loss is higher among older than younger individuals, with over 70% of adults aged 70 and older having hearing loss in at least 1 ear [6]. In the United States (U.S), mild hearing loss affects 23% of the population over the age of 12, with moderate hearing loss more prevalent in those over 80 years [6]. More importantly, approximately 23 million adults with hearing loss in the U.S. do not use hearing aids even though the negative impact of untreated hearing loss has been widely documented [7-9]. The low level of hearing aid adoption is associated with stigma and affordability of hearing aids [8, 15]. Adults with impaired hearing, who do not wear hearing aids demonstrate significantly higher rates of depression, anxiety, and other psychosocial disorders [9,10]. Hearing loss was also associated with increased risk of incident dementia [11-14].

Despite the observed relationship between hearing loss and cognition, surprisingly few studies have investigated the association between the use of hearing aids, cognitive decline, and risk for developing dementia [16-17]. One study found little evidence that hearing aids promoted cognitive function but acknowledged that they may be effective in reducing hearing handicap [18]. Other work showed that hearing aid use was associated with better cognition while controlling for confounding by age, sex, general health, and socioeconomic status [19]. A recent randomized pilot study examined the changes in cognition due to treatment of hearing loss, with some promising results for proximal outcomes (perceived hearing handicap, loneliness) that may mediate a relationship between hearing and cognition [10]. Better understanding of the relationship between use of hearing aids cognitive function, and risk of dementia is of utmost importance since it has the potential to significantly impact public health, as hearing aids represent a minimally invasive, cost effective treatment to mitigate the impact of hearing loss on dementia. In fact, it is postulated that up to 9% of dementia cases could be prevented with proper hearing loss management [14]. Slower conversion from MCI to dementia and progression of dementia following hearing aid use could potentially lead to the reduced incidence of dementia and extended preservation of functional independence in people with dementia.

In this study, we used data from a large referral-based cohort to examine for the first time the effect of the use of hearing aids on the conversion from Mild Cognitive Impairment (MCI) to dementia and risk of death. We tested if the use of hearing aids is independently associated with a decreased risk of incident all-cause dementia diagnosis for MCI patients and reduced risk of death in individuals with dementia. We also examined if the rate of cognitive decline is slower for hearing aid users when compared to those not using hearing aids in participants with both MCI and dementia at baseline.

## 2 Methods

### 2.1 Participants

We conducted a retrospective analysis of the demographic and clinical data obtained from the National Alzheimer’s Coordinating Center (NACC) [20]. The NACC database consists of data from Alzheimer’s Disease Research Centers (ADRCs) supported by the National Institute on Aging (NIA) (grant U01AG016976). Details about the NACC consortium, data collection process, and design and implementation of the NACC database have been reported previously [20,21]. The data set used in our longitudinal investigation was the NACC Uniform Data Set (UDS) [21].

The analytic sample for this study included 2114 participants (age >50) with hearing impairment who had UDS data in the NACC database available between 2005 and 2018 (Fig. 1). All subjects were classified into two groups according to the disease stage. Group 1 included 939 individuals that were diagnosed with MCI at baseline, namely, 497 MCI converters (MCI-c) and 442 MCI non-converters (MCI-nc). Group 2 consisted of 1175 participants that were diagnosed with dementia at baseline: 349 of those died during the follow-up. The 829 dementia participants who did not die during the study follow-up were censored at their last clinical evaluation. Note that only patients that clearly progressed from one stage to another were included in the study. In addition, only active participants who continued to return for annual follow-up visits were taken into account and hence, any subject that missed a scheduled appointment (7%) was discarded from the analysis.

**Fig. 1.**
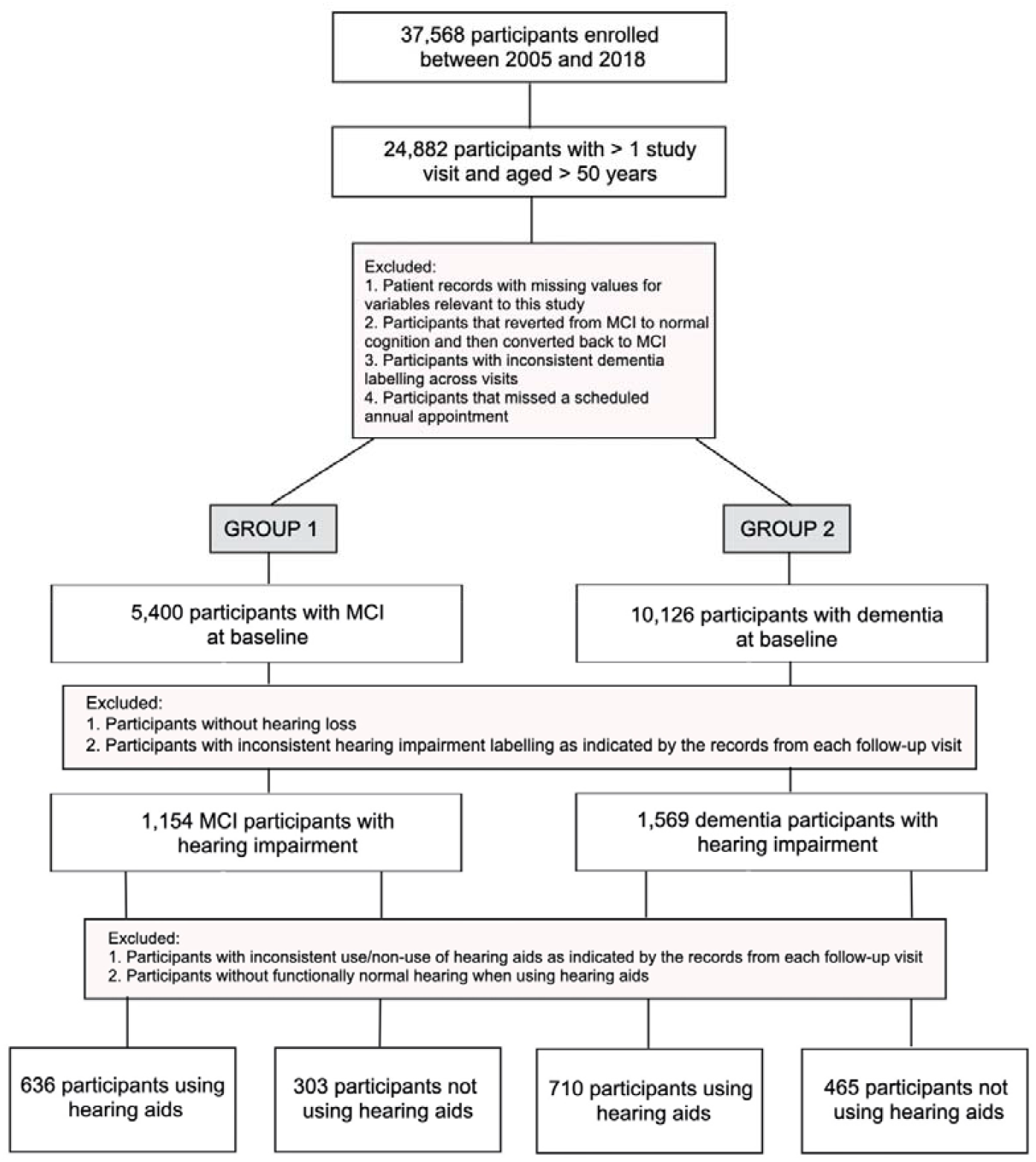
Selection of participants for study inclusion.

### 2.2 Clinical assessment and diagnosis

The incidence of MCI and all-cause dementia was determined based on the clinical diagnosis made by a single clinician or a consensus panel. The clinical diagnosis took into account patient’s medical history, medication use, neuropsychological test performance, and other modifying factors, such as educational and cultural background, and behavioural assessments. In NACC-UDS Version 1 and 2, the procedure of clinical diagnosis of all-cause dementia depended on the diagnostic protocol of the ADRC, but each Center generally adhered to standardized clinical criteria as outlined by the DSM-IV or NINDS-ADRDA guidelines [22,23]. In NACC-UDS Version 3, the coding guidebook criteria for all-cause dementia were modified from the McKhann all-cause dementia criteria (2011) [24]. Diagnoses of MCI were made using the modified Petersen criteria [25].

In addition, we used the CDR® Dementia Staging Instrument Sum of Box (CDRSB) scores to assess a decline due to cognitive changes in six functional domains, namely, memory, orientation, judgment and problem solving, community affairs, home and hobbies, and personal care [26]. CDRSB has a total score range from 0 to 18 points, with higher scores indicating greater cognitive impairment. CDRSB has been commonly used as a reliable tool for assessing dementia severity [27,28].

### 2.3 Hearing assessment

The information on presence of hearing loss and use of hearing aids were extracted from the NACC UDS Physical Evaluation form [29]. Information on hearing loss was collected via self-report using a single hearing screening question: “Without a hearing aid(s), is the subject’s hearing functionally normal?”, which provided possible responses of ‘yes’ or ‘no’ [29]. Participants that responded ‘yes’ were defined as individuals with hearing loss and those that responded ‘no’ were excluded from the analysis. The determination of severity of hearing loss was not a part of a standardized clinical evaluation given to NACC participants. Although participants were allowed to wear hearing aids during the cognitive assessments, it was not reported to NACC whether a participant was wearing a hearing aid during cognitive testing. Missing codes were however entered when ADRCs had a reason to believe that the test was invalid, including if they were aware that the participant was unable to hear properly. All participants with missing codes were excluded from the analysis.

Information on hearing aid use was collected via self-report using a single question: “Does the subject usually wear a hearing aid(s)?”, which provided possible responses of ‘yes’ or ‘no’ [29]. Participants that responded ‘yes’ were classified as hearing aid users and those that responded ‘no’ were classified as non-users of hearing aids. The consistent use (or non-use) of hearing aids was established if the participant answered “yes” or “no” to the above question at every consecutive visit. There was no additional information in the NACC-UDS on the number of hours of daily use of hearing aids, the type of hearing aids used, or the history of hearing aid use before the enrolment in the NACC-UDS.

Accordingly, data from individuals identified with impaired hearing at every annual clinical evaluation, that consistently reported non-use or use of hearing aids as well as functionally normal hearing when wearing hearing aids served as the sample for our study. The subject’s hearing was characterised as functionally normal with a hearing aid if there was no evidence of reduced ability to do everyday activities such as listening to the radio/television or talking with family/friends [29]. This information was based on self-report. On the other hand, the group of participants excluded from the analysis consisted of subjects without hearing impairment or with inconsistent hearing impairment labelling as indicated by the records from each follow-up visit. Participants with inconsistent records relating to the use or non-use of hearing aids or without functionally normal hearing when wearing a hearing aid were also excluded. The baseline characteristics of participants included and excluded from the study are shown in Table A.1. Among 2114 patients with hearing impairment included in the study, 636 subjects in Group 1 and 710 in Group 2 were classified as using hearing aids.

### 2.4 Statistical analysis

Baseline summary statistics are presented as proportions for categorical data and means with standard deviations (SD) for continuous variables. Unadjusted analyses for comparison of demographic and clinical features between individuals with hearing impairment that used and did not use hearing aids were performed with the Fisher’s exact test and unpaired t test [30]. The average annual rate of change in CDRSB score in individuals using and not using hearing aids was compared by applying Mann Whitney U method [30]. The assumption of normality of CDRSB data was tested using Shapiro-Wilk test [30].

A Cox model with censored data was used to study time to incident dementia diagnosis for MCI patients (Group 1) and death for individuals diagnosed with dementia (Group 2) [31]. Censoring was accounted for in the analysis to allow for valid inferences. Ignoring censoring and equating the observed follow-up time of censored subjects with the unobserved total survival time would likely lead to an overestimate of the overall survival probability. The proportional hazards assumption for the Cox regression model fit was tested with the Schoenfeld residuals method and satisfied for Group 1 (*P* = 0.7). The presence of non-proportional hazards was observed in Group 2 (*P* = 0.001), with the proportional hazard assumption violated for age (*P* < 0.001) and CDRSB (*P* = 0.007) variables. Accordingly, we used the Cox proportional hazards regression model to model time to incident dementia for MCI patients (Group 1) and implemented weighted Cox regression accounting for time-varying effects to determine the survival rate of individuals diagnosed with dementia (Group 2) [32]. Weighted Cox regression allowed for providing unbiased estimates of hazard ratios irrespective of proportionality of hazards [32]. Hazard ratios (HR) with 95% confidence intervals (95% CI) were calculated for each group by comparing the hazard rates for individuals with hearing impairment who used and did not use hearing aids. All comparisons were adjusted by age, gender, years of education (measured as the number of years of education completed) and CDRSB score to remove their possible confounding effect. The linearity assumption of the relationship between continuous confounding variables (i.e. age, education, CDRSB) and the log-hazard of the time-to-event outcome was tested using the Box-Tidwell approach and satisfied in both groups (p > 0.05) [33].

For each MCI individual in Group 1 and dementia participant in Group 2, time ‘zero’ was defined as the date of the baseline evaluation. The diagnosis of MCI in Group 1 and dementia in Group 2 referred to the initial event while the endpoint event was considered the conversion to all-cause dementia in Group 1 and the occurrence of death in Group 2 (0 – censored, 1 – uncensored). Survival time was determined by the year. MCI-nc subjects and dementia participants who did not die during the study follow-up were censored at the last clinical assessment.

To avoid the inflation of false-positive findings, the Benjamini-Hochberg false discovery rate (FDR) procedure was used to adjust for multiple hypothesis-testing [34]. False discovery-adjusted *P* values (FDR *P*) less than 0.05 were considered as statistically significant.

### 2.5 Sensitivity analysis

To validate the robustness of the main findings, we performed three different sensitivity analyses: a) propensity score matching to control for measured confounding, b) the analysis for unmeasured confounding to assess the sensitivity of our main conclusions with respect to confounders not included in our study, and c) the inverse probability of treatment weighting method to reduce selection bias within our study population [35,36,37].

Propensity scores were generated for hearing aid status using multivariable logistic regression model and adjusting for baseline covariates, including age, gender, education, and CDRSB. The standardized mean difference between two patient groups i.e., patients with and without hearing aids, was then calculated for each covariate and compared before and after the matching process to determine covariate balance between the two groups [34]. A standardized difference of less than 0.1 was considered negligible in the prevalence of a covariate [34].

Sensitivity analysis for unmeasured confounding was conducted to measure the potential influence an unmeasured covariate might have on the HR estimates of the association between hearing aid use and a) incident dementia in Group 1 and b) death in Group 2 [36]. We considered prevalence rates for the confounder of 5%, 10%, and 20% in the group of hearing aid users and three different values of HR (0.5,2.0,4.0) for the association between the confounder and exposure. We then varied the prevalence of the unmeasured confounder in the group of subjects without hearing aids, from 10% to 30% to determine the extent to which its distribution under these conditions would need to be imbalanced to influence the statistical significance of our findings, i.e., when the upper limit of the 95% CI of HR crosses 1.0.

The inverse probability of treatment weighting method was used to attenuate potential selection bias in the sampling and recruitment of NACC participants. Weights were derived from propensity score modelling of the probability of hearing aid use as a function of measured covariates using the Generalized Boosted Model [37]. A multivariable Cox proportional hazards regression model and weighted Cox regression model were then fitted using derived weights to examine the risk of incident dementia and death for hearing aid users and non-users in Group 1 and 2 respectively.

## 3 Results

### 3.1 Participant Characteristics

Baseline demographic and clinical characteristics of participants by the hearing aid status are presented in Table 1. Statistically significant differences in the use of hearing aids were found between men and women both in Group 1 (*P* = 0.01) and Group 2 (*P* = 0.02), with higher rates of hearing aid use in males. Hearing aid users with baseline MCI were significantly older than MCI individuals not using hearing aids (*P*□<□0.001). Age was comparable for users/non-users of hearing aids in Group 2 (*P* = 0.32). Hearing aid users with dementia had more years of education completed (*P* < 0.001). The CDRSB score was significantly lower in both individuals with baseline MCI and dementia who used hearing aids (*P* = 0.01 and *P* < 0.001 respectively).

**Table 1.** Baseline demographic and clinical characteristics of participants by hearing aid status.

| Characteristic | Group 1 |  |  | Group 2 |  |  |
| --- | --- | --- | --- | --- | --- | --- |
|  | Hearing Aid Status |  |  | Hearing Aid Status |  |  |
|  | Used | Not Used | *P*-value | Used | Not Used | *P*-value |
| Gender, No. (%) |  |  |  |  |  |  |
| Male | 411 (64.6) | 167 (55.1) | 0.01 | 477 (67.2) | 280 (60.2) | 0.02 |
| Female | 225 (35.4) | 136 (44.9) |  | 233 (32.8) | 185 (39.8) |  |
| Age, mean (SD), years | 79.6 (8.9) | 77.3 (8.9) | < 0.001 | 78.2 (7.8) | 77.7 (9.5) | 0.32 |
| Education, mean (SD), years<sup>a</sup> | 16.4 (7.2) | 15.5 (7.6) | 0.07 | 15.4 (3.2) | 14.5 (3.8) | < 0.001 |
| CDRSB score, mean (SD) | 1.4 (1.1) | 1.6 (1.3) | 0.01 | 4.7 (3.0) | 6.5 (4.3) | < 0.001 |

Abbreviation: SD, standard deviation; CDRSB, Clinical Dementia Rating Sum of Boxes
<sup>a</sup> measured as the number of years of education completed

### 3.2 Hearing aid status and risk of incident all-cause dementia in MCI participants

During the study follow-up, 497 MCI subjects in Group 1 developed dementia. The median time to incident dementia was 2 years for non-hearing aid users and 4 years for hearing aid users. The 5-year overall survival rate, which is the percentage of participants that did not convert to dementia 5 year after the baseline MCI diagnosis, was 19% for non-hearing aid users and 33% for individuals using hearing aids.

In the multivariable Cox proportional hazards regression model, the major risk factor for MCI-to-dementia conversion was the CDRSB score (HR 1.39, 95%CI, 1.30-1.48, FDR *P* < 0.001; Table 2), while a significantly reduced risk of dementia was associated with the use of hearing aids (HR 0.73, 95%CI, 0.61-0.89, FDR *P* = 0.004).

**Table 2.**
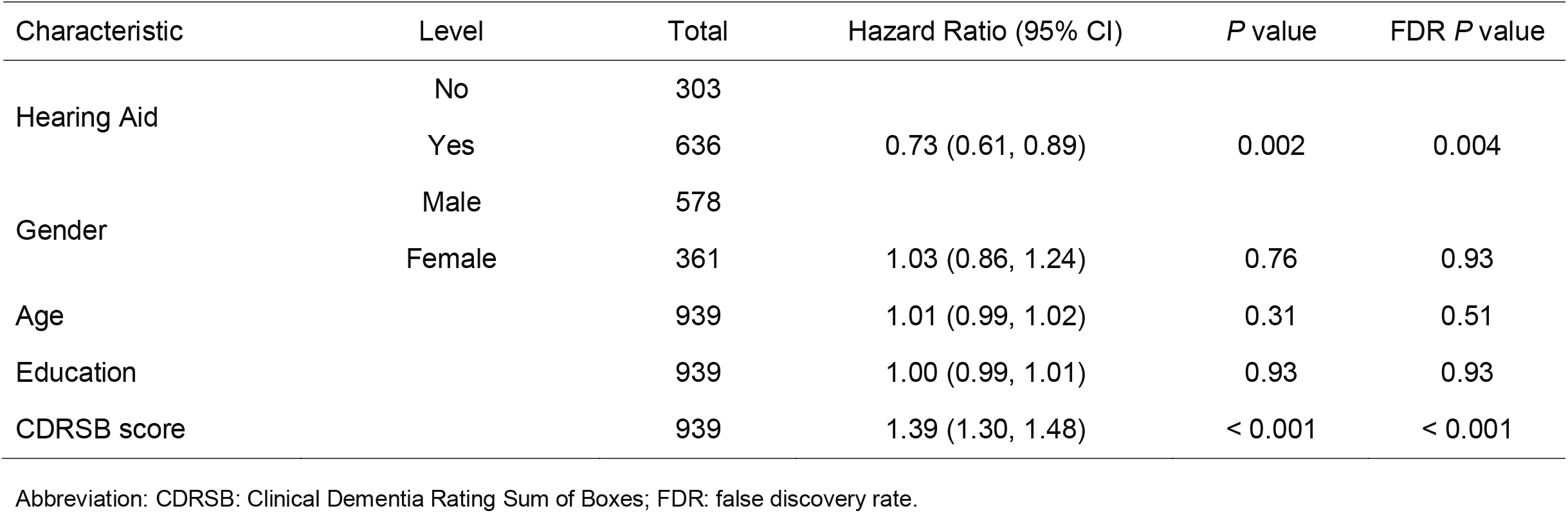
Risk of incident all-cause dementia by hearing aid status after adjustment for age, gender, race, education, and CDRSB score. Hazard ratios with the 95% confidence interval (95%CI) calculated using Cox proportional hazards regression model. FDR: false discovery rate.

| Characteristic | Level | Total | Hazard Ratio (95% CI) | *P* value | FDR *P* value |
| --- | --- | --- | --- | --- | --- |
| Hearing Aid | No | 303 |  |  |  |
|  | Yes | 636 | 0.73 (0.61, 0.89) | 0.002 | 0.004 |
| Gender | Male | 578 |  |  |  |
|  | Female | 361 | 1.03 (0.86, 1.24) | 0.76 | 0.93 |
| Age |  | 939 | 1.01 (0.99, 1.02) | 0.31 | 0.51 |
| Education |  | 939 | 1.00 (0.99, 1.01) | 0.93 | 0.93 |
| CDRSB score |  | 939 | 1.39 (1.30, 1.48) | < 0.001 | < 0.001 |

Abbreviation: CDRSB: Clinical Dementia Rating Sum of Boxes; FDR: false discovery rate.

The observed mean annual (SD) rate of change in CDRSB for non-hearing aid users with MCI was 1.7 (1.95) points per year and significantly higher than the average rate of change for hearing aid users of 1.3 (1.45) CDRSB points per year (*P* = 0.02).

### 3.3 Hearing aid status and mortality risk in participants with dementia

Group 2 consisted of 1175 individuals diagnosed with dementia at baseline: 349 of those died during the follow-up. The median survival time for dementia participants who did not use hearing aids was 6 years and the 5-year overall survival rate was 58%. For hearing aid users, the median survival time was 7 years and the 5-year overall survival rate was 67%.

In the weighted Cox regression model accounting for time-varying effects, the relationship between the use of hearing aids and mortality risk was not statistically significant (HR,□0.98; 95% CI, 0.78-1.24; FDR *P*□=□0.89; Table 3). Higher CDRSB scores were associated with the increased risk of death (HR,□1.08; 95% CI, 1.05-1.11; FDR *P*□<□0.001).

**Table 3.**
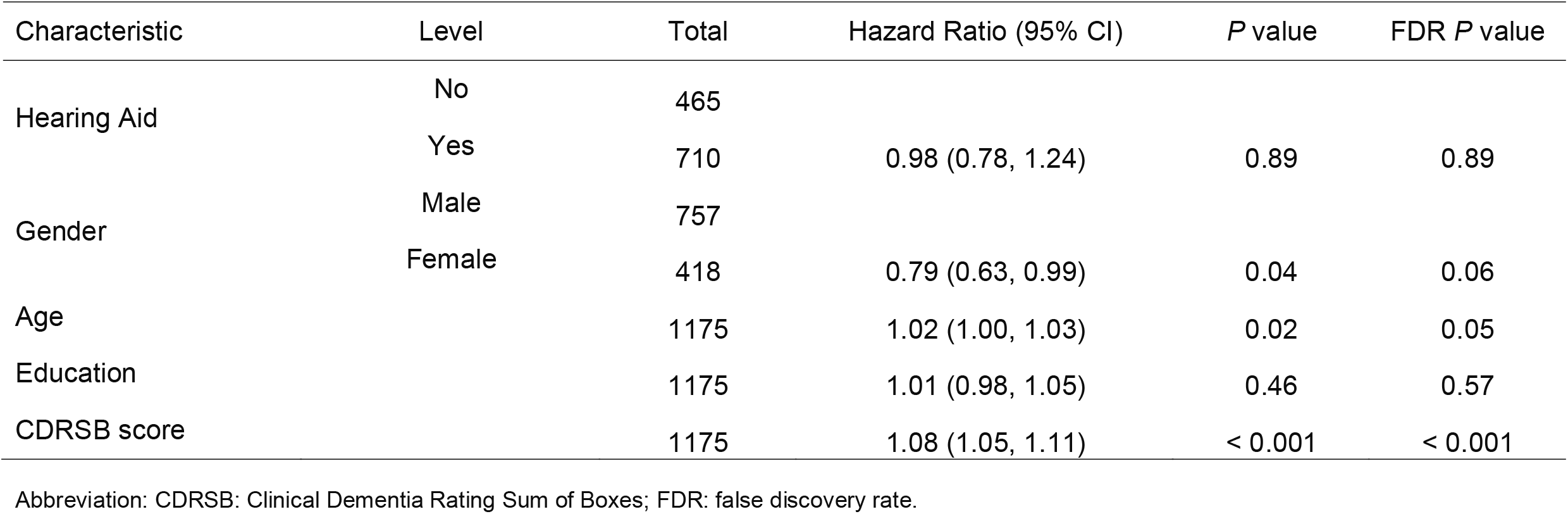
Risk of death by hearing aid status for individuals with dementia after adjustment for age, gender, race, education, and CDRSB score. Hazard ratios with the 95% confidence interval (95% CI) calculated using weighted Cox regression model accounting for time-varying effects. FDR: false discovery rate.

The average (SD) annual rate of change in CDRSB score of 0.96 (1.02) for hearing aid users with dementia was not significantly different from the 0.94 (1.19) point increase in CDRSB score per year for individuals with dementia not using hearing aids (*P* = 0.75).

### 3.4 Sensitivity analysis

The distributions of potential confounders were similar between the hearing aid and non-hearing aid user groups after propensity score matching (standardized difference < 0.1). Again, we found that MCI individuals using hearing aids were at lower risk of developing dementia when compared to non-users of hearing aids (HR 0.67, 95%CI, 0.50-0.80, FDR *P* = 0.01) (Table 4). No link between hearing aid use and risk of death was found for individuals with dementia (Table 4).

**Table 4.** Risk of incident all-cause dementia (Group 1) and risk of death (Group 2) by hearing aid status in the propensity score matched sample.

| Characteristic | Level | Total | Hazard Ratio (95% CI) | P value | FDR P value |
| --- | --- | --- | --- | --- | --- |
| *Group 1* |  |  |  |  |  |
| Hearing Aid | No | 303 |  |  |  |
|  | Yes | 303 | 0.67 (0.50, 0.80) | 0.004 | 0.01 |
| Gender | Male | 427 |  |  |  |
|  | Female | 179 | 0.92 (0.71, 1.19) | 0.52 | 0.65 |
| Age |  | 606 | 1.01 (0.99, 1.02) | 0.48 | 0.65 |
| Education |  | 606 | 0.99 (0.98, 1.01) | 0.9 | 0.91 |
| CDRSB score |  | 606 | 1.37 (1.27, 1.50) | < 0.001 | < 0.001 |
| *Group 2* |  |  |  |  |  |
| Hearing Aid | No | 465 |  |  |  |
|  | Yes | 465 | 0.93 (0.69, 1.27) | 0.66 | 0.66 |
| Gender | Male | 631 |  |  |  |
|  | Female | 299 | 0.91 (0.70, 1.21) | 0.54 | 0.66 |
| Age |  | 930 | 1.01 (0.99, 1.03) | 0.14 | 0.35 |
| Education |  | 930 | 1.02 (0.98, 1.07) | 0.28 | 0.47 |
| CDRSB score |  | 930 | 1.07 (1.03, 1.10) | < 0.001 | 0.002 |

Abbreviation: CDRSB: Clinical Dementia Rating Sum of Boxes; FDR: false discovery rate.

Sensitivity analysis for unmeasured confounding performed for each of two studied groups produced virtually unchanged findings (Table 5). Within Group 1, we observed a lower risk of incident dementia for hearing aid users with the estimated HR for incident dementia in the group of participants with hearing aids below 1 for all considered values of the strength of the confounder-outcome association (HR 0.5, 1.5, 2.0, 4.0), and the prevalence of potential confounder in the group of hearing aid users (5%, 10%, 20%) and non-users (10%, 20%, 30%). No association between hearing aid use and risk of death was found in Group 2.

**Table 5.** Sensitivity analysis for unmeasured confounding

| Prevalence of unmeasured confounder (%) |  | HR adjusted for unmeasured confounder (95% CI)<sup>a</sup> |  |  |  |
| --- | --- | --- | --- | --- | --- |
| Hearing aid used | Hearing aid not used | Unmeasured confounder HR<br>0.5 | Unmeasured confounder HR<br>1.5 | Unmeasured confounder HR<br>2.0 | Unmeasured confounder HR<br>3.0 |
| *Group 1* |  |  |  |  |  |
| 5 | 10 | 0.75 (0.62,0.91) | 0.72 (0.59,0.87) | 0.70 (0.58,0.85) | 0.67 (0.56,0.82) |
|  | 20 | 0.80 (0.66,0.96) | 0.68 (0.57,0.83) | 0.64 (0.53,0.78) | 0.58 (0.48,0.70) |
|  | 30 | 0.84 (0.70,0.97) | 0.65 (0.54,0.79) | 0.59 (0.49,0.72) | 0.51 (0.42,0.61) |
| 10 | 10 | 0.73 (0.61,0.89) | 0.73 (0.61,0.89) | 0.73 (0.61,0.89) | 0.73 (0.61,0.89) |
|  | 20 | 0.78 (0.64,0.94) | 0.70 (0.58,0.85) | 0.67 (0.56,0.82) | 0.63 (0.52,0.76) |
|  | 30 | 0.82 (0.68,0.99) | 0.67 (0.55,0.81) | 0.62 (0.51,0.75) | 0.55 (0.46,0.67) |
| 20 | 10 | 0.70 (0.58,0.84) | 0.77 (0.64,0.93) | 0.80 (0.66,0.97) | 0.84 (0.71,0.99) |
|  | 20 | 0.73 (0.61,0.89) | 0.73 (0.61,0.89) | 0.73 (0.61,0.89) | 0.73 (0.61,0.89) |
|  | 30 | 0.78 (0.64,0.94) | 0.70 (0.58,0.85) | 0.68 (0.56,0.82) | 0.64 (0.53,0.78) |
| *Group 2* |  |  |  |  |  |
| 5 | 10 | 1.03 (0.81,1.31) | 0.98 (0.77,1.25) | 0.96 (0.75,1.22) | 0.92 (0.72,1.17) |
|  | 20 | 1.09 (0.85,1.38) | 0.93 (0.73,1.19) | 0.88 (0.69,1.12) | 0.79 (0.62,1.00) |
|  | 30 | 1.15 (0.90,1.47) | 0.89 (0.70,1.14) | 0.81 (0.63,1.03) | 0.69 (0.54,0.88) |
| 10 | 10 | 1.00 (0.79,1.28) | 1.00 (0.79,1.28) | 1.00 (0.79,1.28) | 1.00 (0.79,1.28) |
|  | 20 | 1.06 (0.83,1.35) | 0.96 (0.75,1.22) | 0.92 (0.72,1.17) | 0.86 (0.67,1.10) |
|  | 30 | 1.12 (0.88,1.43) | 0.91 (0.72,1.17) | 0.85 (0.66,1.08) | 0.75 (0.59,0.96) |
| 20 | 10 | 0.95 (0.74,1.21) | 1.05 (0.82,1.34) | 1.09 (0.86,1.39) | 1.17 (0.92,1.49) |
|  | 20 | 1.00 (0.79,1.28) | 1.00 (0.79,1.28) | 1.00 (0.79,1.28) | 1.00 (0.79,1.28) |
|  | 30 | 1.06 (0.83,1.35) | 0.96 (0.75,1.22) | 0.92 (0.73,1.18) | 0.88 (0.69,1.12) |

Abbreviation: HR, hazard ratio
<sup>a</sup> All models were adjusted for sex, age, education, and CDRSB score

The results of the inverse weighted propensity showed the increased risk of dementia for MCI subjects in Group 1 (HR 0.72, 95%CI, 0.60-0.88, FDR *P* = 0.003) and no statistically significant association between hearing aid use and risk of death for dementia participants in Group 2.

A lack of improvement in hearing, when a hearing aid is used, may be an indicator of central auditory processing issues rather than a faulty device. As such, we performed additional analysis on a group of hearing aid users that included subjects who still experienced auditory difficulties when wearing a hearing aid (Table A.3). In total, 129 participants in Group 1 and 174 participants in Group 2 were identified as those without functionally normal hearing when wearing a hearing aid. A majority of them, 63% in Group 1 and 68% in Group 2, were males. We observed a lower risk of developing dementia in subjects using hearing aids in Group 1 (HR,□0.74; 95% CI, 0.61-0.89; FDR *P*□=□0.003) and no statistically significant relationship between the use of hearing aids and mortality risk in Group 2 (HR,□0.99; 95% CI, 0.80-1.23; FDR *P*□=□0.92).

## 4 Discussion

Despite the prevalence of auditory impairment in dementia, hearing loss is often not diagnosed and not treated even though hearing loss has been shown to be an independent risk factor for poorer cognitive function [38], depression and loneliness [39], and diminished functional status [39]. Several longitudinal studies indicated that individuals with hearing impairment experience substantially higher risk of incident all-cause dementia [12,40-42]. For instance, the study of Lin et al [12] observed a strong relationship between hearing loss and risk of developing dementia. While the authors observed no association between use of hearing aids and reduced risk of dementia, they found a strong link between degree of hearing loss and dementia incidence [12]. In Ray et al [42], the association between cognitive impairment and degree of hearing loss was also observed but only in individuals who did not use hearing aids.

Hypothesized mechanisms explaining the association between hearing impairment and cognitive function included the reallocation of cognitive resources to auditory perceptual processing [43,44,45], cognitive deterioration due to long-term deprivation of auditory input [43,46], a common neurodegenerative process in the aging brain [43], and social isolation caused by both sensory and cognitive loss [45]. In addition, recent findings have suggested that hearing impairment manifested as central auditory dysfunction may be an early marker for dementia [47]. Previous studies concluded that intervention in the form of hearing aids may have a positive effect on cognition [19] and reduce the impact of behavioural and psychological symptoms of dementia [48].

In this study, we investigated the relationship between the use of hearing aids with incidence and progression of dementia. Our results clearly suggest that the use of hearing aids is independently associated with a decreased risk of incident all-cause dementia diagnosis for MCI patients. Statistically significant differences in cumulative survival functions by hearing aid status were found in Group 1, with accelerated cognitive decline, as indicated by change in the CDRSB score in the MCI group. The use of hearing aids was not associated with reduced risk of death in people with dementia. Three different sensitivity analyses confirmed the robustness of our findings.

So far, hearing aid usage has been linked to improvements in cognition as well as psychological, social, and emotional functioning [13]. Amieva *et al*. [49] showed that non-use of hearing aids was associated with faster cognitive decline as indicated by the accelerated rate of change in Mini Mental State Examination score. Yet, no significant difference in cognitive decline was observed between hearing-impaired subjects using hearing aids and healthy individuals [49]. The recent study of Maharani *et al*. [16] adopted a different approach in examining differences in cognitive outcomes of hearing aid use. To prevent potential residual confounding caused by demographic differences between hearing aid users and non-users, the authors analysed rates of cognitive change before and after hearing aid use in the same individuals. They reported a significantly slower decline in episodic memory scores after patients started to use hearing aids.

The potential mechanisms behind the association between the use of hearing aids and cognitive loss, in particular the decreased risk for incident dementia, remain to be determined. Possible explanations include optimized communication and increased social engagement, with resulting lower rates of depression and loneliness, caused by the use of hearing aids and/or changes to the brain associated with the reduced impact of sensory deprivation on brain function [50]. There is also the possibility that facilitated access to auditory information for individuals using hearing aids may result in a reduction in cognitive resources consumed by listening and, hence, lead to improved cognitive ability [50].

We acknowledge that biases in our analysis could arise from multiple sources. First, there is selection bias associated with different recruitment strategies implemented by each ADRC. Those enrolling in ADRC cohorts are not random volunteers and therefore, are not representative of a wider population. Their level of education and income is likely above the national average and approximately 50% of subjects have a family history of dementia. These factors may limit generalizability of our findings. Another methodological limitation of our analysis is its reliance on retrospective data. Even though all ADRCs use standard evaluation procedures, there might be some variation in diagnostic criteria between Centers. The lack of use of consistent diagnostic definitions due to the retrospective design of this study can lead to an increased risk of bias due to potential misclassification of the outcome. Furthermore, bias may arise from the degree of accuracy with which subjects have been classified with respect to their exposure status, i.e. hearing status and hearing aid use status. We minimized this bias by considering only participants with a consistent record of hearing impairment and use/non-use of hearing aids. In this way, we could obtain the true effect of hearing aids on incident and progression of dementia. The inclusion of participants with noisy labels could likely result in an over or underestimation of the effect between exposure (hearing aid use) and outcome (incident dementia or death).

It is also worth highlighting that the proportion of participants wearing hearing aids in the final groups was considerably higher than the prevalence of hearing aid use in the general population [7]. This potential selection bias might have been introduced into the study at the data-gathering stage (the education level and income of NACC volunteers may not be reflective of the general population) or during the process of identifying the study population. In fact, a large number of participants with dementia were excluded from the analysis due to the lack of consistency in reporting hearing difficulty.

Gender, socioeconomic factors and cultural influences all play a role in the use of hearing aids. It may also be the case that unrecorded intrinsic factors that influence use of hearing aids, or lack thereof, play a significant role in the findings presented here. Indeed, severity of cognitive decline may influence acceptance, compliance and correct usage of hearing aids. With incremental changes and decline in cognition, the capacity to comply with the use of hearing aids is likely to significantly diminish [8]. Despite the fact that age, education, gender, and cognitive assessment score were included in the analysis as potential confounders, other unmeasured factors may have impact on the incident and progression of dementia. For instance, hearing aids use in the U.S. is dependent on financial resources as hearing aids are expensive and generally not covered by medical insurance [8]. Other potentially important characteristics not considered in this study due to unavailability of data include type of hearing aid used, hours of daily use, and use of other communicative strategies. Consequently, whether these factors may have a significant effect on time to incident dementia for MCI patients remains unknown and will require further study.

Nevertheless, we implemented measures to account for the potential impact of confounding and selection bias in our study. Propensity score matching was applied to control for measured confounding; the analysis for residual confounding was implemented to assess the sensitivity of our main conclusions with respect to confounders not included in our study; and potential selection bias was addressed via the inverse probability of treatment weighting method. Our results remained robust under different assumptions.

Finally, this study relies on self-reported hearing loss which is far less reliable than audiometric screening. The use of information on hearing impairment via self-report prevents any adjustment for the effect of degree of hearing loss when investigating the impact of hearing aid usage on incidence and progression of dementia. It is also worth noting that even though ADRCs are anticipated to enter missing codes when they have reason to believe that the cognitive test is invalid, including if they are aware that the participant is unable to hear properly, we cannot exclude the possibility that hearing impairment, rather than cognitive function, impacted the ability to complete tasks on cognitive tests in participants with hearing loss who did not have hearing aids. Since verbal instructions that are used during cognitive testing depend significantly on hearing, hearing loss might have contributed to the overestimation of the level of cognitive impairment in some hearing-impaired individuals.

Irrespective of the limitations associated with the present analyses, the fact that significant benefit appears to be derived in delaying conversion of MCI to dementia with hearing aid use warrants further exploration. Properly designed clinical trials will definitively measure the potential benefit of hearing correction in those experiencing hearing loss.

## 5 Conclusion

Slower conversion from MCI to dementia in individuals using hearing aids suggests that effective identification and treatment of hearing loss may reduce the cumulative incidence of dementia. The competing risk of all-cause mortality and dementia among those with MCI should be examined in future work. One of the findings reviewed above suggested that higher-level cognitive processing involving memory in hearing-impaired individuals might be compromised because of mental resources being reallocated to perception and away from storing information. We believe this hypothesis should be further tested to see if the use of hearing aids can make word identification less effortful and thus, allow for freeing resources for higher-level processing that can in turn result in improvement in cognitive function. Furthermore, more research is needed to better understand the relationship between hearing impairment, changes in cognitive ability, and the role of hearing aids in preventing milder forms of cognitive impairment. Such knowledge may provide new and novel insights into prevention of cognitive decline. Most importantly, the magnitude and causality of the effect of hearing loss treatment on cognitive decline and incident dementia can only be established by conducting a well-designed clinical trial.

Public health campaigns are needed to demonstrate the scale and impact of hearing loss and increase awareness regarding effective prevention strategies, consequences of inaction and potential benefits of timely audiological intervention.

## Data Availability

This research was conducted using the National Alzheimer's Coordinating Center database under application 1026.

## Acknowledgements

The NACC database is funded by NIA/NIH Grant U01 AG016976. NACC data are contributed by the NIA-funded ADCs: P30 AG019610 (PI Eric Reiman, MD), P30 AG013846 (PI Neil Kowall, MD), P30 AG062428-01 (PI James Leverenz, MD) P50 AG008702 (PI Scott Small, MD), P50 AG025688 (PI Allan Levey, MD, PhD), P50 AG047266 (PI Todd Golde, MD, PhD), P30 AG010133 (PI Andrew Saykin, PsyD), P50 AG005146 (PI Marilyn Albert, PhD), P30 AG062421-01 (PI Bradley Hyman, MD, PhD), P30 AG062422-01 (PI Ronald Petersen, MD, PhD), P50 AG005138 (PI Mary Sano, PhD), P30 AG008051 (PI Thomas Wisniewski, MD), P30 AG013854 (PI Robert Vassar, PhD), P30 AG008017 (PI Jeffrey Kaye, MD), P30 AG010161 (PI David Bennett, MD), P50 AG047366 (PI Victor Henderson, MD, MS), P30 AG010129 (PI Charles DeCarli, MD), P50 AG016573 (PI Frank LaFerla, PhD), P30 AG062429-01(PI James Brewer, MD, PhD), P50 AG023501 (PI Bruce Miller, MD), P30 AG035982 (PI Russell Swerdlow, MD), P30 AG028383 (PI Linda Van Eldik, PhD), P30 AG053760 (PI Henry Paulson, MD, PhD), P30 AG010124 (PI John Trojanowski, MD, PhD), P50 AG005133 (PI Oscar Lopez, MD), P50 AG005142 (PI Helena Chui, MD), P30 AG012300 (PI Roger Rosenberg, MD), P30 AG049638 (PI Suzanne Craft, PhD), P50 AG005136 (PI Thomas Grabowski, MD), P30 AG062715-01 (PI Sanjay Asthana, MD, FRCP), P50 AG005681 (PI John Morris, MD), P50 AG047270 (PI Stephen Strittmatter, MD, PhD)

## Ethics approval and consent to participate

The National Alzheimer’s Coordinating Center Uniform Data Set (NACC-UDS) is approved by the University of Washington Institutional Review Board and participants provided informed consent at the ADRC where they completed their study visits. The approval was received to use the NACC-UDS data in the present work (application reference: 1026)

## Supplementary material

**Table A.1.** Baseline demographic and clinical characteristics of participants included and excluded from the study.

|  | Group 1 |  |  | Group 2 |  |  |
| --- | --- | --- | --- | --- | --- | --- |
|  | MCI at baseline |  |  | Dementia at baseline |  |  |
|  | Excluded | Included |  | Excluded | Included |  |
| Total | 4461 | 939 |  | 8951 | 1175 |  |
| Gender, No. (%) |  |  |  |  |  |  |
| Male | 2055 (46.1) | 578 (61.6) | ] < 0.001 | 4259 (47.6) | 757 (64.4) | ] < 0.001 |
| Female | 2406 (53.9) | 361 (38.4) |  | 4692 (52.4) | 418 (35.6) |  |
| Age, mean (SD), years | 74.7 (9.0) | 78.9 (8.0) | < 0.001 | 73.4 (9.6) | 78.0 (8.5) | < 0.001 |
| Education, mean (SD), years<sup>a</sup> | 15.5 (5.9) | 16.1 (7.4) | 0.03 | 15.3 (8.3) | 15.1 (3.5) | 0.12 |
| CDRSB score, mean (SD) | 1.4 (1.1) | 1.5 (1.2) | 0.01 | 5.7 (3.9) | 5.4 (3.7) | 0.01 |

Abbreviation: SD, standard deviation; CDRSB, Clinical Dementia Rating Sum of Boxes
<sup>a</sup> measured as the number of years of education completed

**Table A.2.**
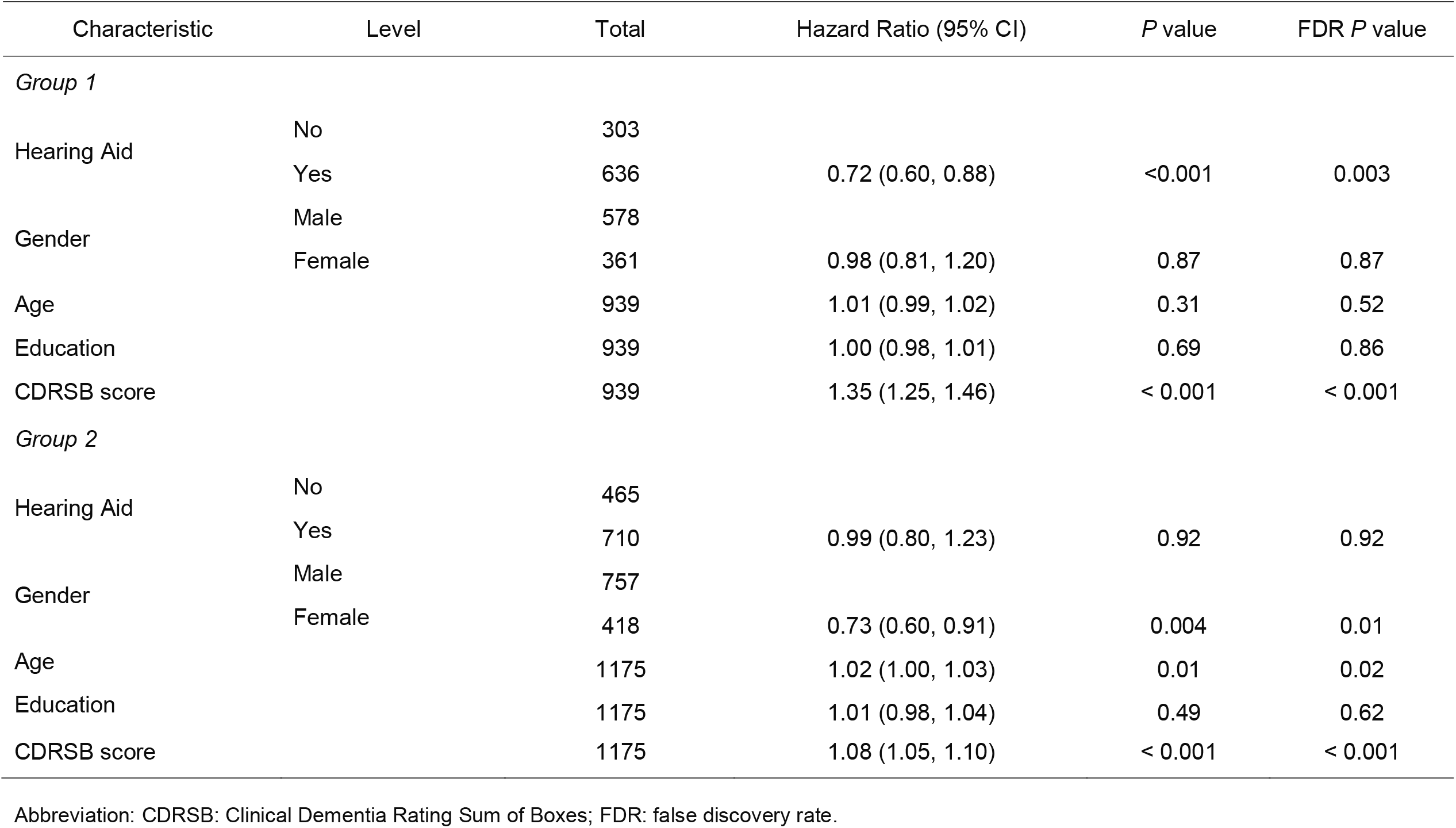
Sensitivity analysis based on the inverse probability weighting

| Characteristic | Level | Total | Hazard Ratio (95% CI) | P value | FDR P value |
| --- | --- | --- | --- | --- | --- |
| *Group 1* |  |  |  |  |  |
| Hearing Aid | No | 303 |  |  |  |
|  | Yes | 636 | 0.72 (0.60, 0.88) | <0.001 | 0.003 |
| Gender | Male | 578 |  |  |  |
|  | Female | 361 | 0.98 (0.81, 1.20) | 0.87 | 0.87 |
| Age |  | 939 | 1.01 (0.99, 1.02) | 0.31 | 0.52 |
| Education |  | 939 | 1.00 (0.98, 1.01) | 0.69 | 0.86 |
| CDRSB score |  | 939 | 1.35 (1.25, 1.46) | < 0.001 | < 0.001 |
| *Group 2* |  |  |  |  |  |
| Hearing Aid | No | 465 |  |  |  |
|  | Yes | 710 | 0.99 (0.80, 1.23) | 0.92 | 0.92 |
| Gender | Male | 757 |  |  |  |
|  | Female | 418 | 0.73 (0.60, 0.91) | 0.004 | 0.01 |
| Age |  | 1175 | 1.02 (1.00, 1.03) | 0.01 | 0.02 |
| Education |  | 1175 | 1.01 (0.98, 1.04) | 0.49 | 0.62 |
| CDRSB score |  | 1175 | 1.08 (1.05, 1.10) | < 0.001 | < 0.001 |

Abbreviation: CDRSB: Clinical Dementia Rating Sum of Boxes; FDR: false discovery rate.

**Table A.3.** Risk of incident all-cause dementia (Group 1) and risk of death (Group 2) by hearing aid status. The group of hearing aid users consisted of subjects with functionally normal hearing with a device and those that exhibited reduced ability to do everyday activities such as listening to the radio/television or talking with family/friends when wearing a hearing aid

| Characteristic | Level | Total | Hazard Ratio (95% CI) | P value | FDR P value |
| --- | --- | --- | --- | --- | --- |
| *Group 1* |  |  |  |  |  |
| Hearing Aid | No | 303 |  |  |  |
|  | Yes | 765 | 0.74 (0.61, 0.89) | 0.001 | 0.003 |
| Gender | Male | 659 |  |  |  |
|  | Female | 409 | 1.05 (0.89, 1.25) | 0.55 | 0.69 |
| Age |  | 1068 | 1.01 (0.99, 1.02) | 0.32 | 0.54 |
| Education |  | 1068 | 1.00 (0.99, 1.01) | 0.82 | 0.82 |
| CDRSB score |  | 1068 | 1.40 (1.32, 1.49) | < 0.001 | < 0.001 |
| *Group 2* |  |  |  |  |  |
| Hearing Aid | No | 465 |  |  |  |
|  | Yes | 884 | 0.99 (0.80, 1.23) | 0.92 | 0.89 |
| Gender | Male | 876 |  |  |  |
|  | Female | 473 | 0.73 (0.60, 0.91) | 0.004 | 0.01 |
| Age |  | 1349 | 1.02 (1.00, 1.03) | 0.01 | 0.02 |
| Education |  | 1349 | 1.01 (0.98, 1.04) | 0.49 | 0.61 |
| CDRSB score |  | 1349 | 1.07 (1.05, 1.10) | < 0.001 | < 0.001 |

Abbreviation: CDRSB: Clinical Dementia Rating Sum of Boxes; FDR: false discovery rate.

